# Prevalence of polymyxin resistant bacterial strains in India: a systematic review and meta-analysis

**DOI:** 10.1101/2023.10.04.23296553

**Authors:** Sambit K. Dwibedy, Indira Padhy, Aditya K. Panda, Saswat S. Mohapatra

## Abstract

**Introduction:** Polymyxins, the cationic lipopeptide antibiotics, are the last line of therapeutics against the MDR Gram-negative bacterial (GNB) pathogens. Unfortunately, the rising cases of polymyxin-resistant strains from across the globe have adversely impacted their utility. While the molecular mechanisms responsible for developing polymyxin resistance (Pol^R^) are largely understood, the prevalence of Pol^R^ strains in India has not been investigated systematically. The current study was undertaken to primarily determine the prevalence of Pol^R^ strains in India. Moreover, the extent of the spread of mobile colistin resistance (*mcr*) genes among the GNB strains in India was also determined.

**Method:** A systematic search for articles using the relevant inclusion and exclusion criteria was performed in the applicable databases for the period January 2015 to December 2023. The included 41 studies were subjected to a meta-analysis using the Comprehensive Meta-Analysis software (V.4). Publication biases were assessed using funnel plots and Egger’s regression analysis.

**Result:** Considering a total of 41 studies including 24,589 bacterial isolates the present meta-analysis found the rate of Pol^R^ bacteria in India to be at 15.0% (95% CI: 11.2 to 19.8). Among the Indian States, Tamil Nadu topped with the highest prevalence of Pol^R^ at 28.3%. Investigating the contribution of the *mcr* genes, it was observed that among the Pol^R^ strains, 8.4% (95% CI: 4.8 to 14.3) were *mcr* positive.

**Conclusion:** The study determined the prevalence of Pol^R^ strains in India at 15.0% which is higher than that of the global average at 10%. The study also determined that 8.4% of the Pol^R^ strains carried the *mcr* genes. The *mcr*-positive strains reported from India could be an underestimation of the actual numbers due to the non-inclusion of *mcr* screening in many previous studies. This study provides insight into the state of the Pol^R^ situation in India, which may be useful to develop a monitoring strategy to contain the spread of such strains and preserve the efficacy of the polymyxins.

## INTRODUCTION

The significant use of antibiotics in healthcare, and agriculture, and their indiscriminate prophylactic consumption has driven the rise of antibiotic-resistant bacterial pathogens. Infections caused by antibiotic-resistant bacteria are increasingly becoming fatal due to the non-availability of effective antibiotics against them. As per an estimate, drug-resistant pathogens caused 4.95 million deaths in 2019,^1^ which is predicted to rise to 10 million deaths per annum by 2050.^2^ Considering the significant economic consequences of rising drug-resistant infections, urgent mitigation measures are warranted.

Antibiotic-resistant bacteria belonging to the Gram-negative type pose a formidable challenge to treatment as increasing reports of resistance to third and fourth generations of cephalosporins are reported globally.^3^ In this unprecedented scenario, polymyxins, a group of cationic lipopeptide antibiotics, remain the last option for treatment.^4,5^ Polymyxin was initially isolated in 1947 from a soil bacterium *Paenibacillus polymyxa*^6^ and approved for clinical use in 1959 against Gram-negative bacteria (GNB). Due to their nephrotoxic and neurotoxic properties, polymyxins were prohibited for clinical use in the 1970 s^7,8^ and were consequently substituted by aminoglycosides, quinolones, and β-lactams. However, with the rising infections caused by MDR GNB such as *Klebsiella pneumoniae*, *Acinetobacter baumannii*, and *Pseudomonas aeruginosa*, and failure of the frontline antibiotics, polymyxins have re-emerged albeit with some modifications in the chemical structure as the last-resort antibiotics against them.^4,9^

Polymyxins are of five types (polymyxin A to polymyxin E), of which polymyxin B and polymyxin E (colistin) are approved for clinical use.^10^ Structurally, polymyxins are cationic decapeptides consisting of a central cyclic heptapeptide with a tripeptide side-chain acylated at the N-terminus by a fatty acid tail.^11,12^ Polymyxins primarily target the negatively charged outer membrane (OM) of GNB, where they disrupt the lipopolysaccharide (LPS) structure causing membrane damage and cell death.^5,13,14^

The extensive use of polymyxins in the preceding decade in both healthcare and animal feeds has triggered the emergence of polymyxin-resistant (Pol^R^) bacterial strains. Pol^R^ bacterial species such as *K. pneumoniae*,^15–17^ *P. aeruginosa*,^4,18^ *A. baumannii*,^19^ *E. coli*,^20^ *Shigella sonnei*,^21^ *Salmonella enterica*,^22^ *Enterobacter aerogenes* and *Enterobacter cloacae*^23^ have been reported from across the globe adversely affecting the healthcare sector. Moreover, the detection of Pol^R^ GNB in both clinical and environmental samples has made the situation concerning.

The primary mechanism responsible for Pol^R^ development is the OM remodelling or modification in GNB making it increasingly positive [by the addition of 4-amino-4-deoxy L-arabinose (L-Ara4N) or phosphoethanolamine (pEtN)], leading to the repulsion of cationic polymyxins.^4,13,24^ The OM remodelling is generally carried out by bacterial two-component signal transduction systems (TCS), of which PhoPQ and PmrAB are the prominent ones.^13,24^ Moreover, bacterial species such as *Serratia marcescens*, *Proteus mirabilis*, *Providencia rettgeri*, *Vibrio cholerae*, *Campylobacter spp.*, *Legionella spp.*, *Morganella morganii*, *Burkholderia spp.*, *Edwardsiella spp.*, *Aeromonas hydrophila*, *Acinetobacter junii*, *Neisseria spp*. etc. are intrinsically resistant to polymyxins due to the permanent modification of their OM.^4,24,25^

Chromosome-encoded polymyxin resistance though is the prevalent mechanism, bacteria can acquire polymyxin resistance via the plasmid-borne mobile colistin resistance (*mcr*) gene. In this context, the first detection of *mcr* plasmids in 2015 from *E. coli* and *K. pneumoniae* strains in China is significant.^20^ The *mcr* gene encodes a pEtN transferase enzyme that catalyses the addition of a pEtN residue to lipid-A moiety which alters the net charge of the LPS. To date, ten different classes of *mcr* genes (*mcr-1* to *mcr-10*) including several variants have been discovered among many GNB pathogens.^26,27^ Plasmids carrying *mcr* genes have been reported from across the globe showing distinct geographical distribution.^26,28^ As the potential of rapid dissemination of plasmid-borne *mcr* genes is significantly high, it is pertinent to conduct periodic surveillance to monitor their distribution in the environment and clinic.

In India, *mcr* genes have been reported in several studies conducted using clinical samples,^29^ hospital sewage,^30^ urban sewage water,^31^ and food samples.^32^ However, a comprehensive analysis of the distribution of *mcr* genes across India is lacking. It is important to highlight that the increasing number of carbapenem-resistant pathogenic bacteria has made India one of the top users of polymyxins in the hospital setting. Furthermore, the use of colistin as a growth promoter in poultry farms has triggered the spread of Pol^R^ GNB in the environment. Though the Govt. of India has banned the use of colistin in farm animals since 2019, the impact of their extensive use before 2019 is yet to be assessed.

Studies reporting the prevalence of Pol^R^ strains in India are very limited and the reports of *mcr*-positive bacterial strains are also rare. The current study employing a systematic review and meta-analysis was undertaken to gain an insight into the prevalence of Pol^R^ bacterial strains across India from both clinical and environmental sources. Moreover, the study also attempted to estimate the *mcr*-positive bacterial strains in India with a particular emphasis on their geographical distribution in the Indian sub-continent.

## MATERIALS & METHODS

Using published articles, the present systematic review and meta-analysis was conducted to determine the prevalence and spread of Pol^R^ GNB from clinical and environmental sources in India and the contribution of *mcr* genes in gaining the Pol^R^ phenotype. The study was undertaken as per the guidelines of Systematic Reviews and Meta-Analyses (PRISMA)^33^ (Fig. 1) (Supplementary Table 3).

**Figure. 1.**
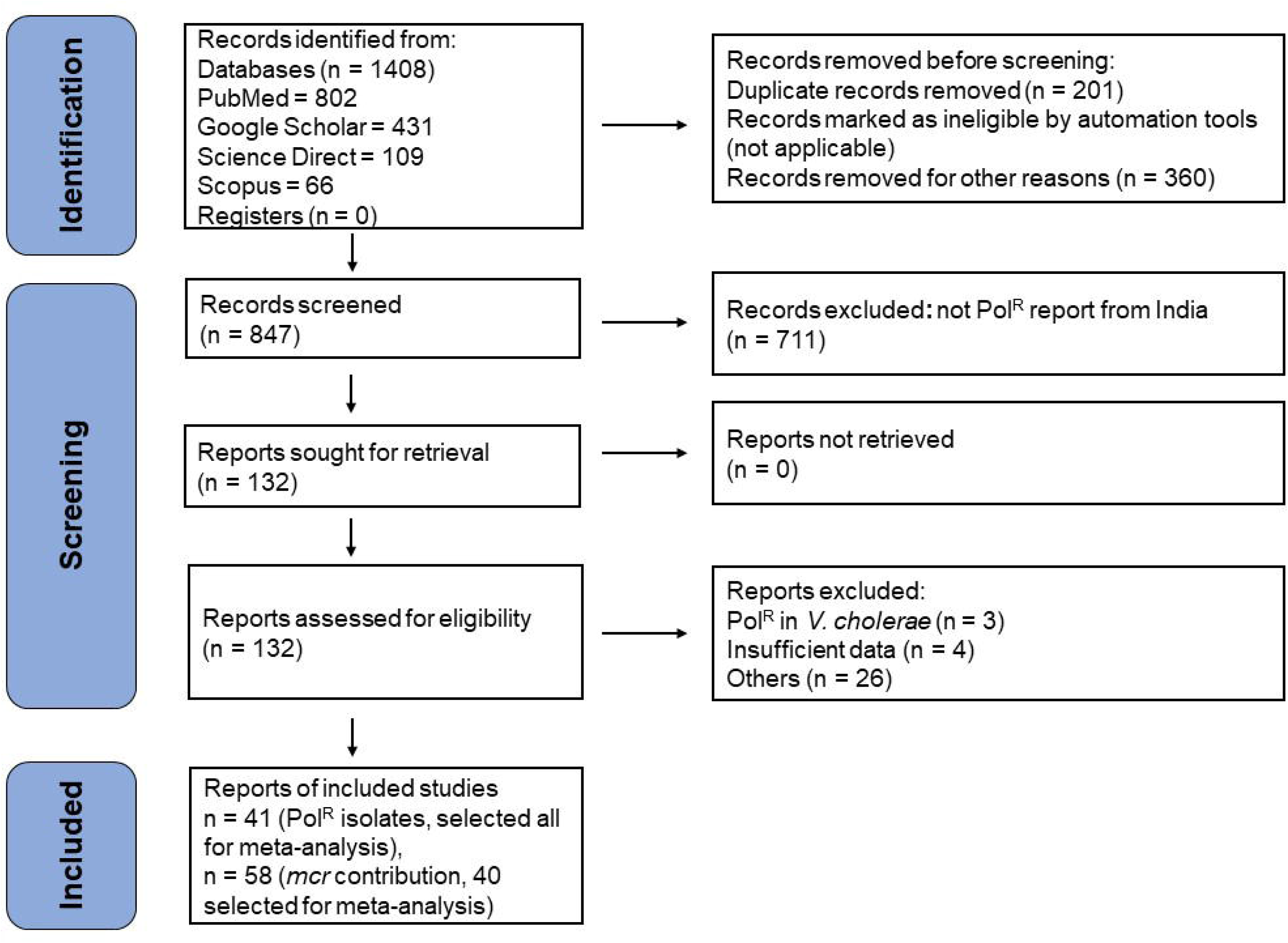
The PRISMA flow diagram for the selection, inclusion, and exclusion of studies.

### Data collection

The databases such as PubMed, Google Scholar, Science Direct, and Scopus were extensively searched using the keywords “polymyxin resistance”, “colistin resistance”, “mobile colistin resistance gene” and “India”. Original studies reporting the presence of Pol^R^ bacterial strains as well as *mcr*-positive strains in India, published between January 2015 to December 2023 were included in this study. As the first report of *mcr*-positive bacterial strains came in 2015,^20^ we selected the timeline from 2015 to 2023 for the meta-analysis to determine the prevalence of both Pol^R^ and *mcr*-positive bacterial strains in India.

### Inclusion criteria

As the primary objective of the study was to determine the prevalence of Pol^R^ bacterial strains in India, all studies reporting such strains irrespective of their origin were considered. Studies involving surveillance of hospital samples, sewage samples, food materials, etc. were analysed. A bacterial isolate was considered Pol^R^ if the MIC to colistin was > 2 mg/L determined using the broth microdilution (BMD) method^34,35^, whereas a strain was considered *mcr*-positive if a particular *mcr* class or any of its variants was amplified by PCR. For the meta-analysis, the following inclusion criteria were followed: 1) the samples used in the studies must hail from India irrespective of their source of isolation, 2) data on resistance to polymyxin B and/or colistin must be available, 3) BMD method must have been used to determine Pol^R^ of the bacterial isolates, 4) the article must mention the total sample size and the number of Pol^R^ strains isolated or the prevalence frequency, 5) the sample size should not be smaller than five for Pol^R^ and one for *mcr* gene positive, and 6) the article must have been published in English.

### Exclusion criteria

The following exclusion criteria were strictly followed during the analysis: 1) studies with no Pol^R^ isolate were excluded; 2) studies conducted outside India were not considered; 3) studies that employed resistance detection methods other than BMD were excluded; 4) studies that were not published in English, were not peer-reviewed, and were published before 2015 were excluded.

### Data extraction

Data were extracted from relevant articles that were qualified based on the study’s inclusion and exclusion criteria (Fig. 1). Each report was individually screened and investigated for eligibility by two different authors (SKD and IP). Following a thorough review, the following information was extracted and summarised for each study: first author name, publication year, State, or UT (Union Territories) of occurrence, total sample size, the total number of Pol^R^ isolates, reported Pol^R^ bacteria, resistance detection method, genes involved in resistance development, and *mcr* gene screening status. For further analysis, the data were tabulated in a Microsoft Excel sheet.

### Subgrouping of data

The research articles with Pol^R^ reports were classified into two groups based on criteria such as sample size (more than five) and the involvement of *mcr* genes in the development of Pol^R^. Studies reporting a specific number of samples containing both Pol^R^ and susceptible isolates and a particular number of Pol^R^ bacteria as events were placed in the first group. This group was further subdivided based on two criteria: geographical location (the States or UTs from where they were reported) and the source of their isolation (clinical or environmental). In this classification system, those States were included that have at least four different reports on Pol^R^ isolates. In the second group, those reports were included where *mcr* gene screening status was mentioned clearly in the study.

### Quality assessment

The quality of all eligible reports was determined by the Newcastle–Ottawa Scale (NOS) and only the high-quality reports were considered for the present meta-analysis. According to the NOS guidelines, qualitative scores are given as stars, and a total score of five stars or more for a study is considered a high-quality report.

### Meta-analysis

The meta-analysis was performed using the relevant studies and reports to determine the prevalence of Pol^R^ bacteria in India, and the contribution of *mcr* genes in Pol^R^ phenotype. All statistical analyses were performed using the Comprehensive Meta-Analysis (CMA) software (version 4.0; Biostat Inc. USA). The generated data were visualized using forest plots. Publication bias in the included studies was assessed using funnel plots with visual analysis^36^ and Egger regression analysis.^37^ The heterogeneity of the studies was examined using the Cochrane Q statistics^38^ and I^2^ statistics.^39^ Based on the outcome of the heterogeneity analysis, a fixed-effect model (homogenous) or random-effect model (heterogeneous) was used for the meta-analysis.

The combined event rate and 95% confidence interval were calculated to determine the frequency of Pol^R^ isolates in India and the contribution of *mcr* to polymyxin resistance. In addition, sensitivity analysis was performed to explore the robustness of the meta-analysis by excluding one study at a time, performing the meta-analysis, and comparing it with the results of the parent analysis.

## RESULTS

### Literature review and screened results

Overall, 1408 articles were obtained by searching different databases with suitable keywords (PubMed = 802, Google Scholar = 431, Science Direct = 109, Scopus = 66). The article selection approach is presented in the PRISMA flow chart (Fig. 1). A total of 41 eligible articles reporting Pol^R^ (Supplementary Table 1) were included in the meta-analysis. For determining the contribution of *mcr* genes in Pol^R^, 58 studies were considered of which 40 studies were found to be eligible for the meta-analysis (Supplementary Table 2) based on the stipulated inclusion and exclusion criteria.

### Publication bias

Visual analysis of the funnel plot and Egger’s regression analysis showed the absence of publication bias in the analysis of the incidence of Pol^R^ bacteria in India (intercept: 0.37, 95% CI: −2.73 to 3.46, p = 0.81), as well as in the analysis of the role of the *mcr* gene in Pol^R^ (intercept: −1.3, 95% CI: −2.69 to 0.08, p = 0.065) (Table 1) (Supplementary Fig. 1).

**Table 1.** Statistics to test the publication bias and heterogeneity.

| Study Name<br>(Incidence of Pol<sup>R</sup><br>bacteria) | Egger's Regression Analysis |  |  | Heterogeneity Analysis |  |  | Model<br>Used |
| --- | --- | --- | --- | --- | --- | --- | --- |
|  | Intercept | 95%<br>Confidence<br>Interval | P Value<br>(2 tailed) | Q value | P-Heterogeneity | I<sup>2</sup> (%) |  |
| India | 0.37 | -2.73 to 3.46 | 0.81 | 1515.89 | 0.00 | 97.36 | Random |
| Delhi | -6.25 | -21.90 to 9.4 | 0.33 | 167.66 | 0.00 | 97.02 | Random |
| Odisha | 2.05 | -5.52 to 9.62 | 0.49 | 83.95 | 0.00 | 94.04 | Random |
| Tamil Nadu | 2.62 | -6.92 to 12.17 | 0.53 | 121.59 | 0.00 | 94.24 | Random |
| Uttar Pradesh | 0.72 | -13.49 to 14.93 | 0.9 | 102.7 | 0.00 | 94.16 | Random |
| India (Clinical) | -0.15 | -3.2 to 2.9 | 0.92 | 1164.99 | 0.00 | 96.99 | Random |
| India (Environmental) | -4.66 | -28.12 to 18.8 | 0.57 | 999.46 | 0.00 | 97.1 | Random |
| *mcr* gene in Pol<sup>R</sup> | -1.3 | -2.69 to 0.08 | 0.06 | 129.71 | 0.00 | 69.93 | Random |

Furthermore, after the categorization of all studies according to the geographical locations (States) from which they were reported, the funnel plot and Egger’s regression analysis revealed no significant biases while analysing the incidence of Pol^R^ bacteria in states such as Delhi, Odisha, Tamil Nadu, and Uttar Pradesh (Table 1) (Supplementary Fig. 2).

### Heterogeneity analysis

Cochrane Q and I^2^ measurements demonstrated significant heterogeneity among the included studies. Hence, a random-effects model was used for all meta-analyses (Table 1).

### Prevalence of Pol^R^ bacteria in India

After analysing 41 reports, accounting for 24,589 bacterial isolates tested, the present meta-analysis found that the rate of Pol^R^ bacteria in India was 15.0% (95% CI: 11.2 to 19.8) (Fig. 2A). Out of the 28 states and eight UTs in India, 12 states and 3 UTs have reported the presence of Pol^R^ bacterial strains. Interestingly, 16 states and 5 UTs in India have not reported the presence of Pol^R^ strains. The overall spread of the Pol^R^ GNB strains in the Indian subcontinent is shown in Fig. 2B. Among all the states that have reported Pol^R^, Tamil Nadu, Uttar Pradesh, and Odisha are the top three with the rate of Pol^R^ at 28.3%, 16.3%, and 15.9%, respectively, which is higher than the overall prevalence of Pol^R^ in the Indian subcontinent (Fig. 3). Moreover, among the other states in India Pol^R^ prevalence in Delhi was found to be quite high at 13.7%, though less than the national average (Fig. 3).

**Figure. 2.**
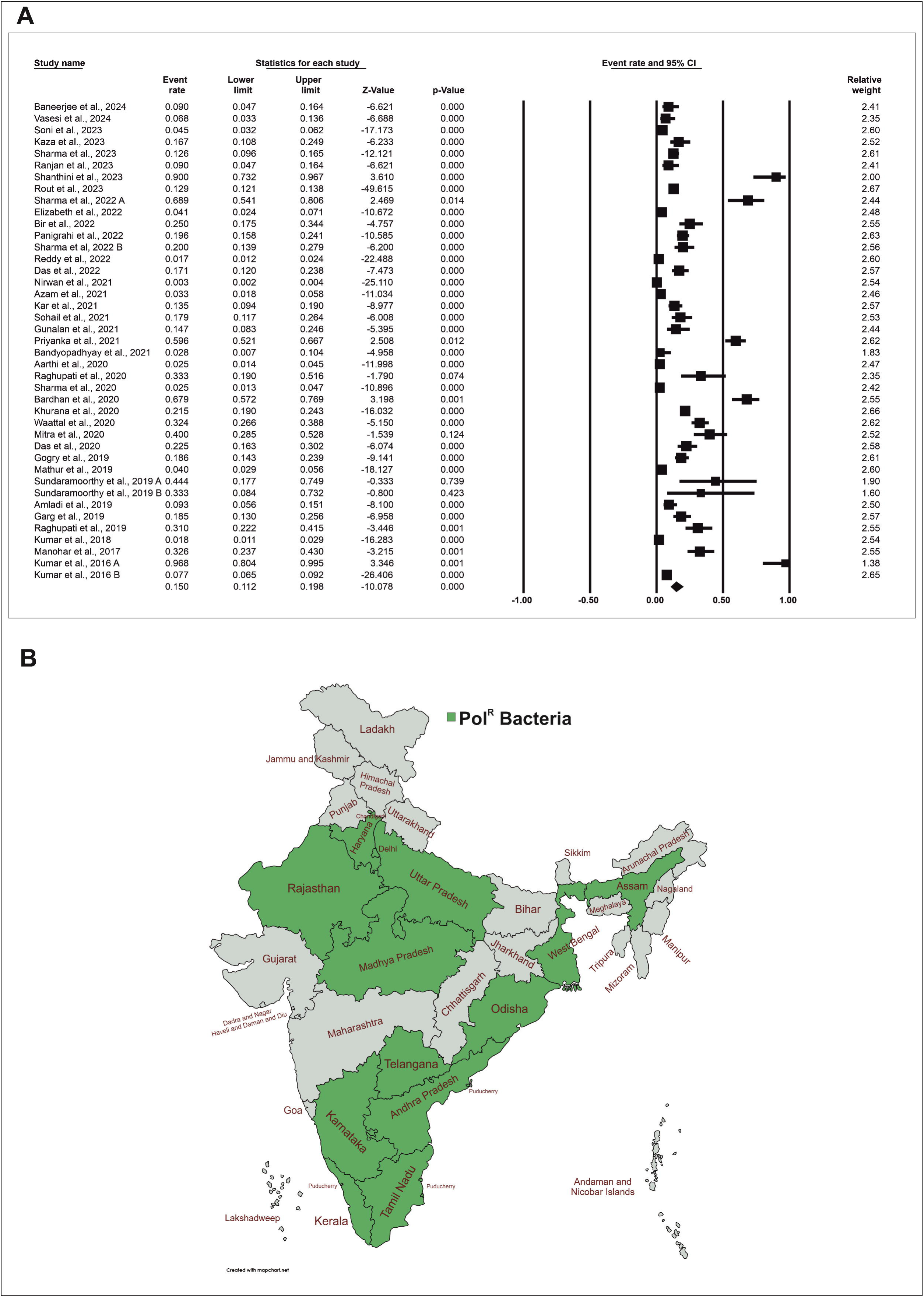
(A) Forest plots demonstrating the event rate of Pol^R^ bacteria in India. The Comprehensive Meta-Analysis software V4 (Biostat Inc. USA) was used for the calculation of the event rate and 95% confidence interval. (B) Distribution of Pol^R^ bacteria across different States and Union Territories (UTs) of India from 41 studies. States and UTs in India that have reported Pol^R^ strains are shaded in green.

**Figure. 3.**
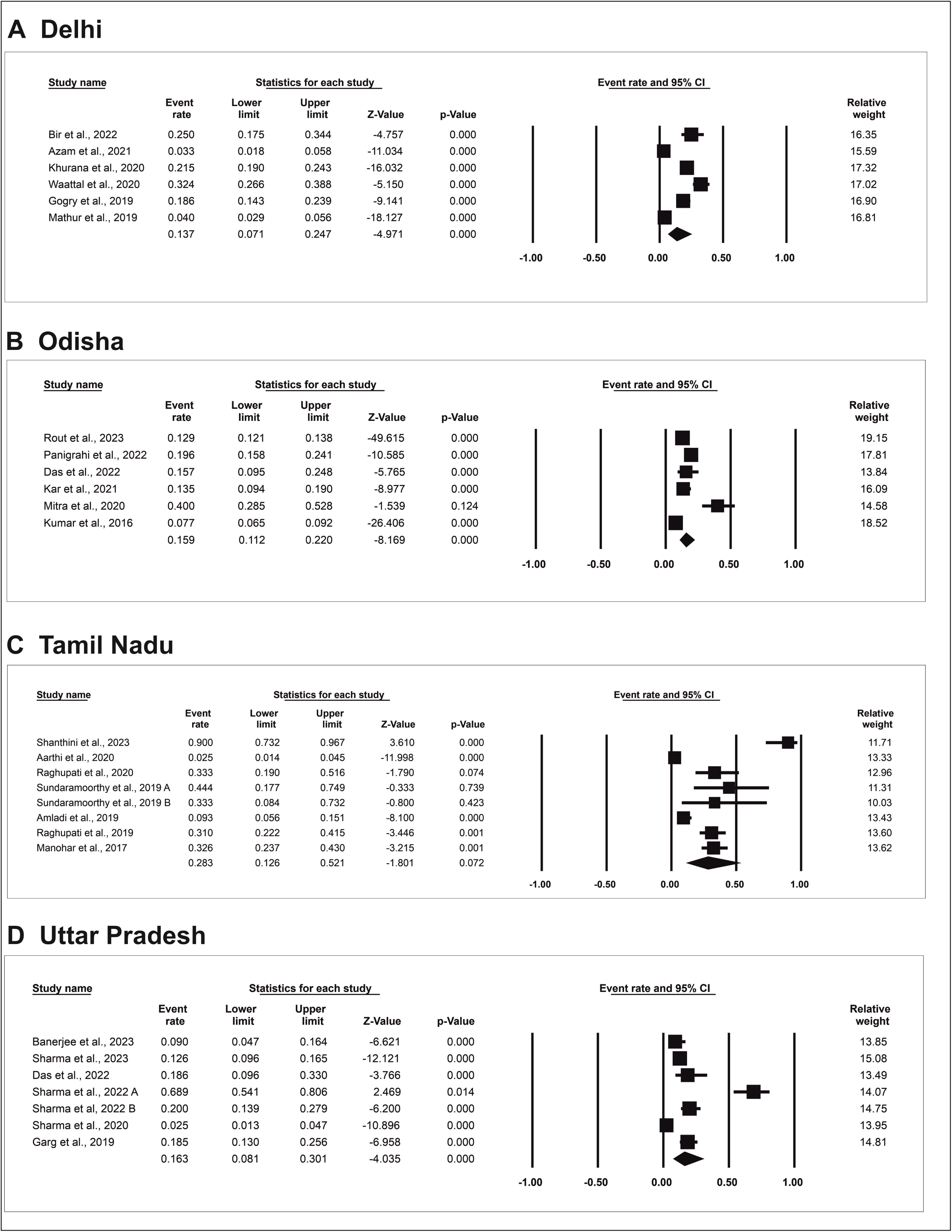
Forest plots representing the event rate of Pol^R^ bacteria in different states of India (A) Delhi, (B) Odisha, (C) Tamil Nadu, and (D) Uttar Pradesh.

A breakdown of the Pol^R^ GNB strains by the source of their isolation revealed that 89% (1875 isolates) of them were from clinical sources, whereas the remaining 11% (227 isolates) were from the environmental samples (Fig. 4A). Bacterial strains isolated in hospitals from various samples such as stool, pus, urine, wound etc. were considered as clinical isolates though further sub-categorization was not performed in the current study. However, among the environmental sources, water (46%), vegetables (45%), poultry products (8%), and dairy farms (1%) were the major contributors for Pol^R^ strains (Fig. 4A). Employing meta-analysis on 36 eligible clinical reports, the rate of Pol^R^ was found to be 13.5% (95% CI: 10.0 to 17.9) (Fig. 4B), whereas a similar analysis using 5 environmental reports the Pol^R^ rate was found to be 27.6% (95% CI: 11.0 to 54.2) (Fig. 4C).

**Figure. 4.**
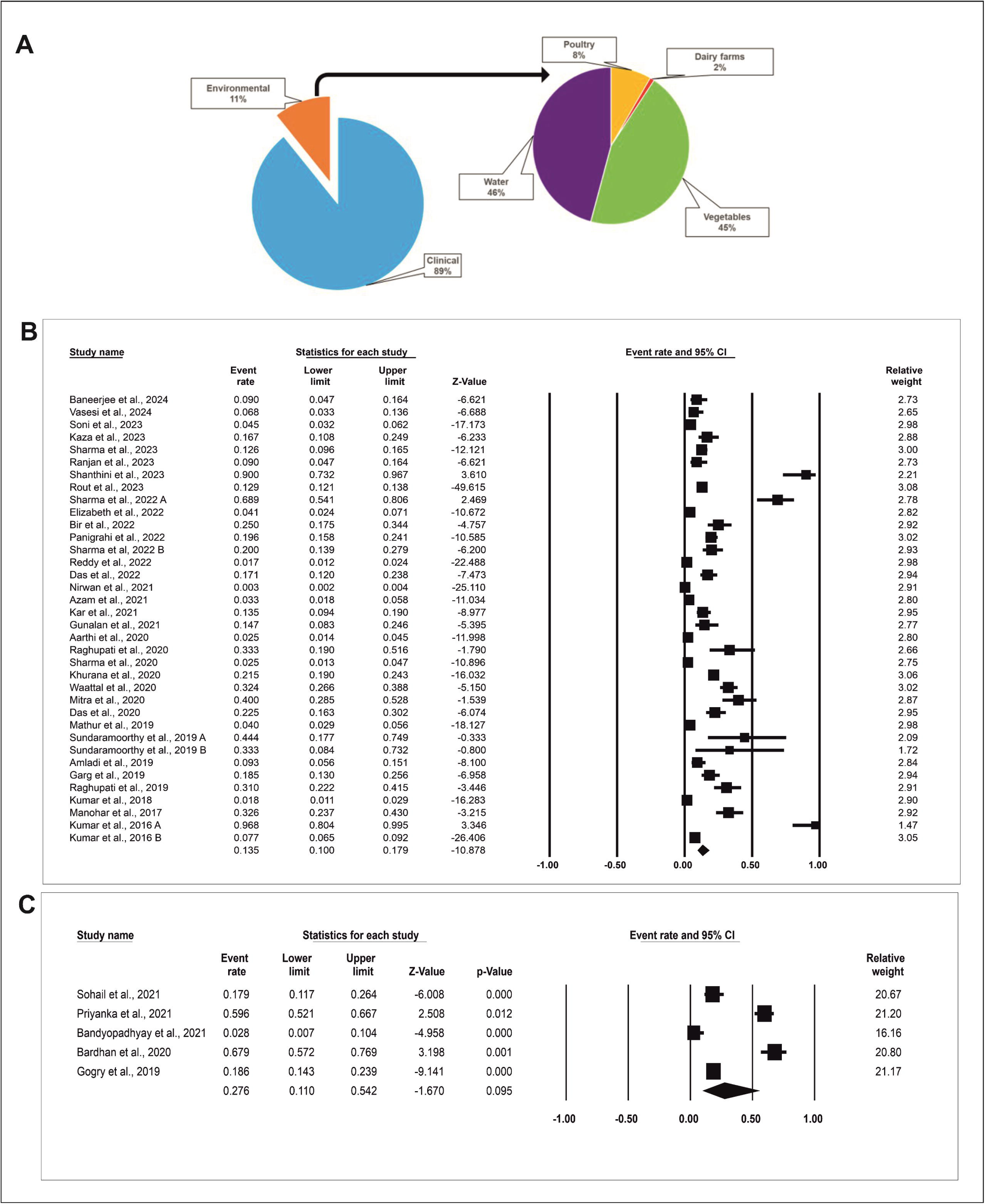
(A) Pie chart showing the prevalence of Pol^R^ bacteria in India based on the source of their isolation. (B) Forest plots revealing the event rate of Pol^R^ bacteria in clinical isolates in India. (C) Forest plots demonstrating the event rate of Pol^R^ bacteria in environmental isolates in India.

### Role of *mcr* gene in the development of Pol^R^

To understand the prevalence of *mcr* genes in Pol^R^ bacterial isolates in India in comparison with other possible mechanisms, 40 eligible reports with positive *mcr* gene screening status and a sample size of more than one (Pol^R^ isolate) criterion were considered for the analysis (Supplementary Table 2). Out of the 1606 bacterial isolates tested for the presence of *mcr* genes by PCR, 76 strains were found to be positive. The meta-analysis result showed that the contribution of the *mcr* gene to the rise in polymyxin resistance was 8.4% (95% CI: 4.8-14.3) in India (Fig. 5A).

**Figure. 5.**
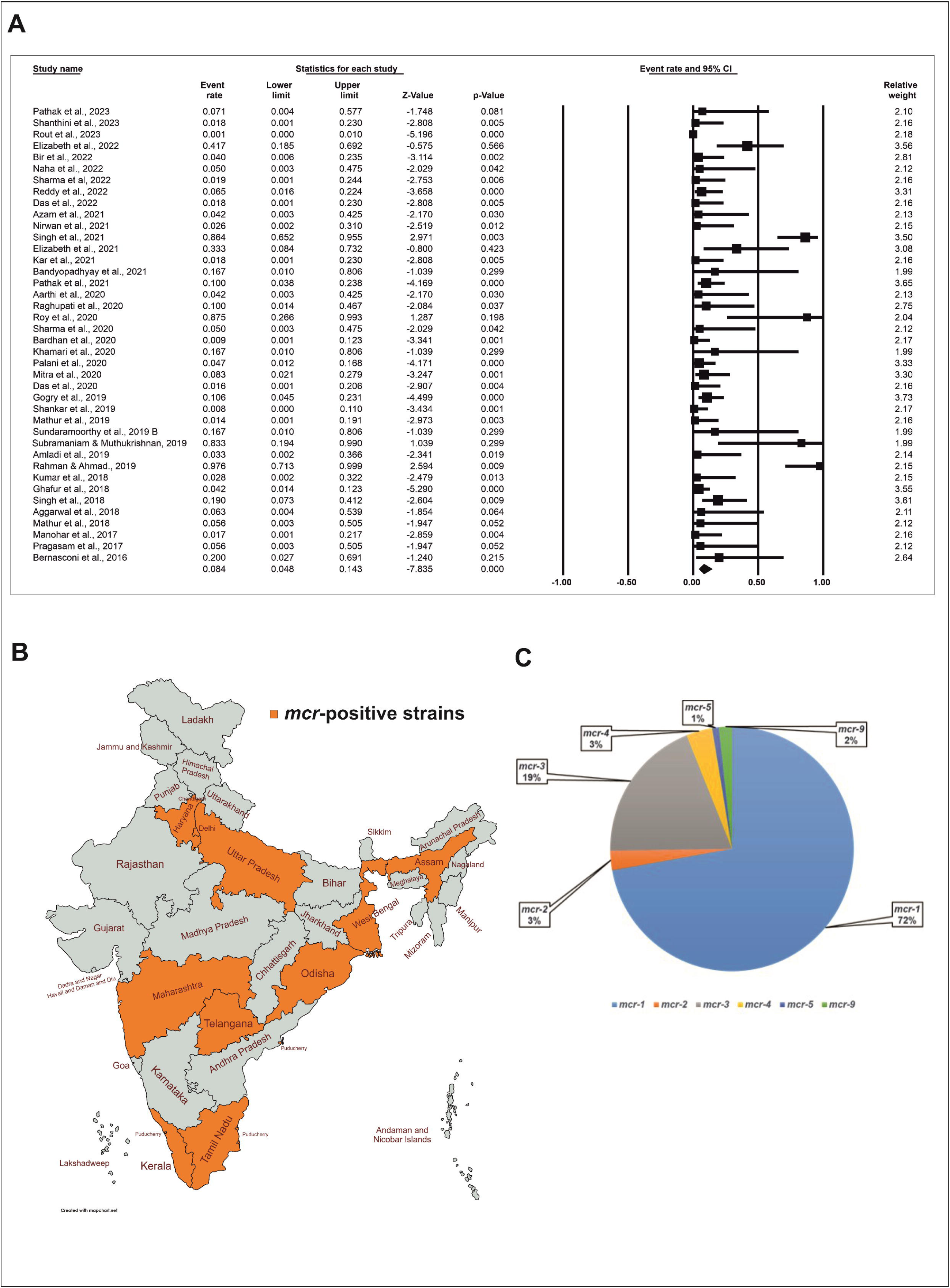
(A) Forest plots demonstrating the contribution of the *mcr* genes in the development of polymyxin resistance in India. (B) Spread of *mcr* genes across different States and UTs of India. States and UTs that have reported *mcr*-positive strains are shaded in orange. (C) Pie chart showing the prevalence of reported classes of *mcr* genes in India.

Analysing the State wise detection of the *mcr* genes, nine states were found to have reported the presence of *mcr* genes (Fig. 5B), among which Uttar Pradesh has the highest number of reports at 48 followed by Tamil Nadu and Assam with 31 and 15 reports respectively. Analysing the prevalence of particular classes of *mcr* genes, it was revealed that in India, so far six classes of *mcr* genes namely *mcr-1*, *mcr-2*, *mcr-3*, *mcr-4*, *mcr-5*, and *mcr-9* have been detected (Table 2). Among the *mcr* classes, the present analysis identified the *mcr-1* to be the predominant one and found in 72% of the isolates, whereas the frequencies of *mcr-2*, *mcr-3*, *mcr-4*, *mcr-5*, and *mcr-9* were 3%, 19%, 3%, 1%, and 2%, respectively (Fig. 5C).

**Table 2.**
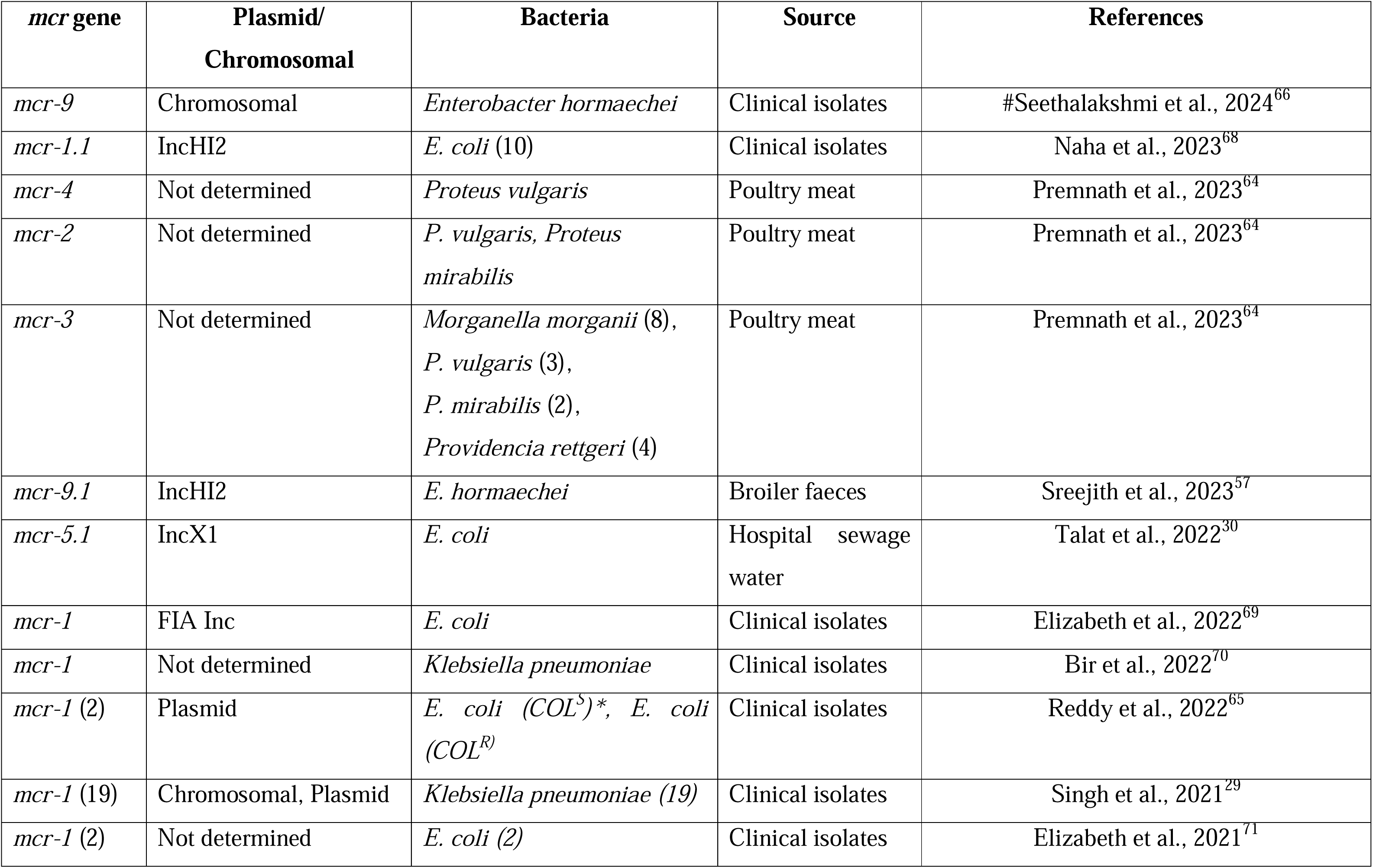
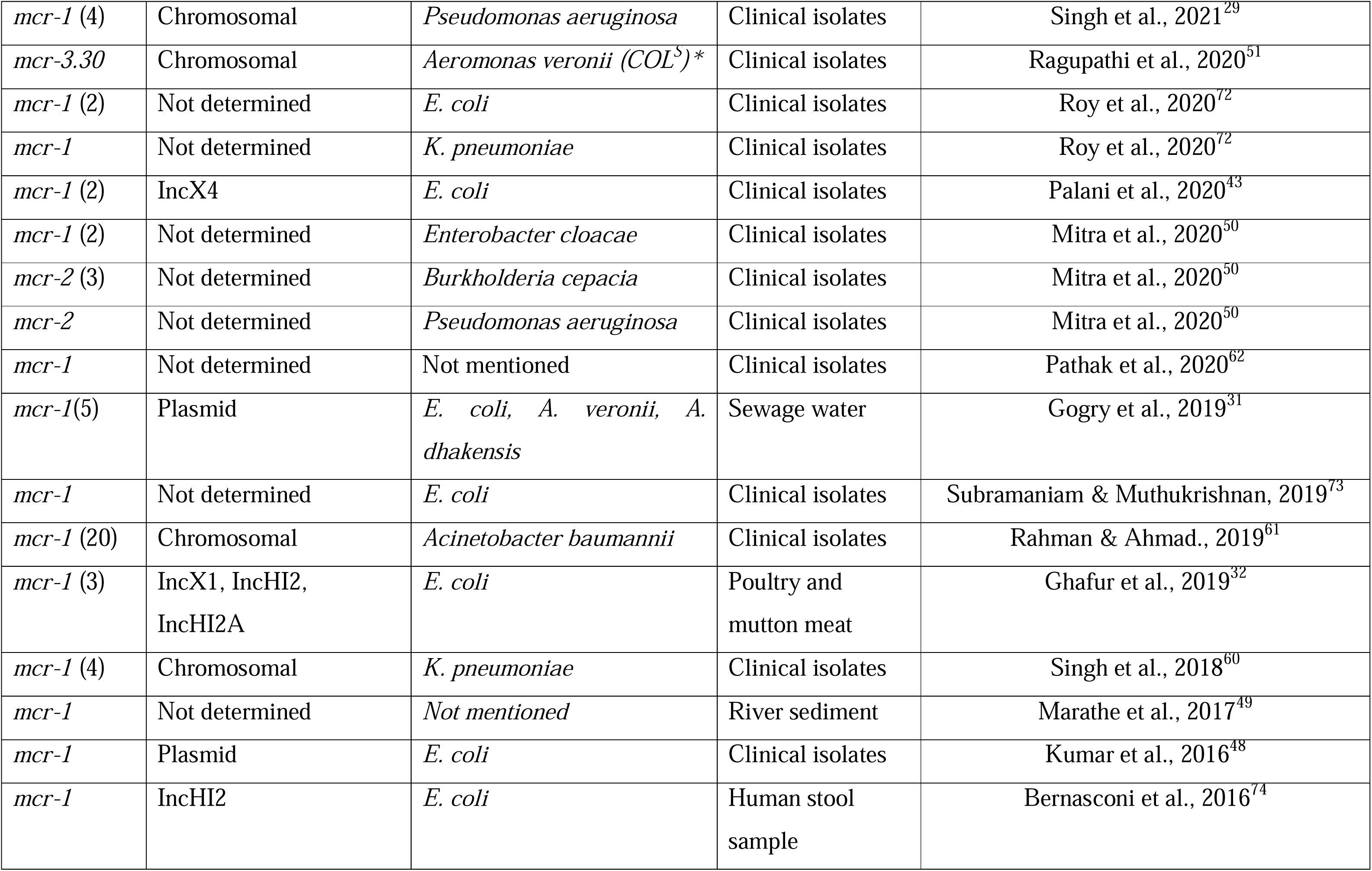
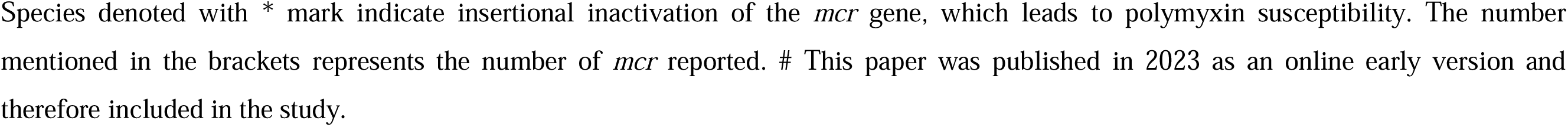
Detailed information on the *mcr* gene reported from India.

### Sensitivity analysis

Sensitivity analysis was performed to measure the robustness of the meta-analysis by excluding a single study each time and relating the results to the parental meta-analysis. As illustrated in Supplementary Fig. 3, no significant deviation was observed in the sensitivity analysis compared with the original meta-analysis results for studying the incidence of Pol^R^ bacteria in India. Furthermore, the meta-analysis on the contribution of the *mcr* genes to Pol^R^ also revealed a robust observation, as shown in Supplementary Fig. 4.

## DISCUSSION

The current meta-analysis systematically evaluated the prevalence of Pol^R^ bacteria in India, which was higher than the global average. In this study, we found that 15.0% of GNB were Pol^R^ (Fig. 2), whereas the global average remained at 10%.^40^ A previous study has put the rate of Pol^R^ among clinical Enterobacteriaceae isolates in India at 13.8%.^41^

The present study revealed that GNB resistant to polymyxins is spread all over India, and States like Tamil Nadu, Uttar Pradesh, and Odisha have a higher prevalence of Pol^R^ strains than the national average (Fig. 2B, Fig. 3). The rising incidence of Pol^R^ in India could be attributed to their indiscriminate use in the healthcare sector, in poultry farms as growth promoters, and the rapid dissemination of *mcr* genes via horizontal gene transfer. Even though India has banned the use of colistin in animal feeds since 2019, its implications on the spread of Pol^R^ may be revealed a few years down the line. Interestingly, there are no reports of Pol^R^ in 16 states and 5 UTs in India, which may be due to the lack of surveillance in those States and UTs (Fig. 2B).

Although most of the reports used in this study indicated Pol^R^ rates under 20%, there are a few that have reported significantly higher Pol^R^ rates.^42–47^ These high numbers are possibly due to the different methodologies adopted for determining polymyxin resistance. In India, various methods such as the Kirby-Bauer disk diffusion test, BMD, agar dilution, Etest, VITEK-2, gradient diffusion, colistin broth disc elution, rapid polymyxin Nordmann Poirel Test, Mikrolatest kit, BD Phoenix M50, MicroScan WalkAway 96 Plus, Micronaut-S and Modified Stokes DDT, have been used. However, as per the recommendation of the CLSI and EUCAST, BMD is the only approved method for determining polymyxin susceptibility,^34,35^ and therefore in this study, the polymyxin resistance reports determined using the BMD method have only been included.

Though the primary goal of this meta-analysis was to find the overall prevalence of Pol^R^ bacterial strains in India, our study revealed that 13.5% of the clinical isolates and 27.6% of the environmental isolates from India are resistant to polymyxins (Fig. 4). However, there were only five eligible studies that were used to analyse the environmental samples (Fig. 4C). Therefore, a larger study might be needed to confirm the observed high rate of resistance among the environmental isolates.

This study revealed that in 8.4% of cases, the *mcr* gene was responsible for the Pol^R^ phenotype (Fig. 5A). In India, after the first detection of the *mcr* gene in a clinical *E. coli* isolate,^48^ several reports of *mcr* occurrence have been published.^30,32,49–51^ To date, six classes of *mcr* genes have been reported in India: *mcr-1*, *mcr-2*, *mcr-3*, *mcr-4*, *mcr-5*, and *mcr-9* (Table 2). Of the ten different *mcr* classes described till date, *mcr-1* was the predominant variant responsible for driving polymyxin resistance in India. In the current study, the frequency of *mcr-1* was 72% (Fig. 5C). Our analysis corroborates the recent studies that have determined the predominance of *mcr*-1 gene in comparison to other *mcr* classes in the spread of colistin resistance.^52–54^ Considering that *mcr-1* was the first plasmid-borne colistin resistance gene to be detected^20^ it has spread across the globe in the preceding decade or so. Moreover, various retrospective surveillance for *mcr* genes have revealed their presence in samples dating back to 1980s highlighting their presence before the widespread use of colistin in the animal feeds, which has only supplemented its global spread.^26,55^

Unlike the distribution of Pol^R^ GNB, the spread of *mcr* has been limited to nine states and two UTs in India (Fig. 5B). Among these states, Uttar Pradesh was found to be the hotspot for the spread of *mcr* variants, with 48 reports, followed by Tamil Nadu and Assam with 31 and 15 *mcr* positive reports, respectively (Supplementary Table 2). However, several reports on Pol^R^ have not provided sufficient information on *mcr* gene screening status. The limited screening for *mcr* genes among these studies may be responsible for the low rate of detection and reporting of *mcr* genes in India.

As mentioned previously, *mcr* genes are largely plasmid-borne, and various types of plasmids are involved in carrying *mcr* genes in GNB.^26,56^ In India, different *mcr* variants were found to be linked with plasmids including IncHI2, InCHI2A, IncX1, IncX4, and FIA Inc.^32,43,57^ The role of these plasmids conferring resistance to carbapenems, cephalosporins, and fosfomycin has also been established previously.^58,59^ Although plasmids are the primary mobile genetic elements through which *mcr* genes disseminate among bacterial species, their chromosomal integration has also been reported in different bacterial species in India.^60–63^ The major advantage of chromosomal integration of *mcr* genes could be their efficient vertical transmission in the absence of polymyxin selection. Some intrinsically resistant bacteria also harbour the *mcr* genes and can potentially spread them to the polymyxin-susceptible species in the environment.^64^ Moreover, few studies have reported the presence of the *mcr-1, mcr-3*, and *mcr-9* genes in bacterial species that are susceptible to polymyxin.^51,65^ This is primarily due to the insertional inactivation of *mcr-1* and *mcr-3* genes.^51,65^ However, in the case of *mcr-9* positive *Enterobacter hormaechei* strain, *qseBC* TCS genes were absent downstream^66^ which has been previously described to induce the colistin resistance phenotype by upregulating the *mcr-9* gene expression in response to the subinhibitory concentration of colistin.^67^

This meta-analysis has certain limitations. First, the current study included articles available in the English language only, as a result, publications in other languages have been excluded. Second, we searched reports from databases including PubMed, Google Scholar, Scopus, and Science Direct, so may have missed publications indexed in other databases. Third, as certain GNB species are intrinsically resistant to polymyxins, reports on polymyxin resistance from the environmental strains may have been biased toward such species.

## CONCLUSION

The present study found that the rate of Pol^R^ bacterial strains in India was 15.0%, which is higher than the global average. Pol^R^ was determined among 13.5% and 27.6% of the bacterial isolates from the clinical and environmental sources respectively. While Pol^R^ strains have been reported from most of the Indian States and UTs, there are still regions where adequate data are not available, warranting monitoring and surveillance. This study further found that in 8.4% of Pol^R^ strains, the phenotype was determined by *mcr* genes carried on plasmids. This may be grossly underestimated due to the lack of *mcr* surveillance data in many published articles. As polymyxins are one of the last option antibiotics available against GNB pathogens, efforts to increase their clinical longevity is paramount. However, the increasing incidence of polymyxin resistance in clinics and the environment has the potential to limit antibiotic treatment options. Therefore, along with developing better versions of polymyxins and/or their derivatives, it is also important to devise strategies for monitoring the frequency of Pol^R^ bacterial species and, the prevalence of *mcr* genes in them. Sustained measures need to be in place to contain further spread of the polymyxin resistance phenotype in the near future.

## Data Availability

All data produced in the present study are available upon reasonable request to the authors.

## Author Contributions

Conceptualization: SKD, AKP, and SSM; Data extraction and analysis: SKD, IP, AKP, and SSM; Data validation: SKD and IP; Manuscript writing and reviewing: SKD, AKP, and SSM. All authors agreed to the submitted version.

## Funding

This work is supported by the funds received from the Science and Technology Department, Govt. of Odisha (Grant no. ST-BT-MISC-0005-2023-2463/ST, dt. 23-05-2023).

Infrastructure support from the “Centre of Excellence on Bioprospecting of Ethno-pharmaceuticals of Southern Odisha (CoE-BESO)” to the Dept. of Biotechnology, Berhampur University is gratefully acknowledged. Indira Padhy is a recipient of the “Biju Patnaik Research Fellowship (BPRF)” from the Science and Technology Department, Govt. of Odisha.

## Transparency Declarations

The authors declare that the research was conducted in the absence of any commercial or financial relationships that could be construed as a potential conflict of interest.

## Notes

### Competing Interest Statement

The authors have declared no competing interest.

### Summary of Updates

The result section has been revised considering new studies. Figures have been updated.

