## Supplementary Information for "Prevalence of polymyxin resistant bacterial strains in India: a systematic review and meta-analysis"

Saswat S. Mohapatra, Ph.D.

Assistant Professor, Department of Biotechnology, Berhampur University, Bhanja Bihar, Berhampur- 760007.

**Supplementary Table 1**. Reports on polymyxin resistance ($Pol^R$) in India.

| Sl. No. | Author | Sample Size | $Pol^R$ Isolates | State / UT | Resistance Detection Method | *mcr* Screening |
|---|---|---|---|---|---|---|
| 1 | #Banerjee et al., 2024[1] | 100 | 9 | Uttar Pradesh | BMD | NO |
| 4 | Vasesi et al., 2024[2] | 103 | 7 | Chandigarh | BMD | NO |
| 2 | Soni et al., 2023[3] | 736 | 33 | Madhya Pradesh | BMD | *mcr 1-3* |
| 3 | Kaza et al., 2023[4] | 108 | 18 | Chandigarh | BMD | YES |
| 5 | Sharma et al., 2023[5] | 356 | 45 | Uttar Pradesh | KB DDT, BMD, ADM | NO |
| 6 | Ranjan et al., 2023[6] | 100 | 9 | Andhra Pradesh | Gradient diffusion, BMD, E-test | NO |
| 7 | Shanthini et al., 2023[7] | 30 | 27 | Tamil Nadu | BMD | *mcr 1-5* |
| 8 | Rout et al., 2023[8] | 6013 | 778 | Odisha | VITEK-2, BMD, CBDE | *mcr 1-5* |
| 9 | Sharma et al., 2022[9] | 45 | 31 | Uttar Pradesh | KB DDT, BMD | NO |
| 10 | Elizabeth et al., 2022[10] | 291 | 12 | Assam | BMD, RPNP | YES |
| 11 | Bir et al., 2022[11] | 100 | 25 | Delhi | BMD, VITEK-2, RPNP, E-test | *mcr 1-5* |
| 12 | Panigrahi et al., 2022[12] | 357 | 70 | Odisha | VITEK-2, BMD | NO |
| 13 | Sharma et al, 2022[13] | 125 | 25 | Uttar Pradesh | BMD, CBDE | *mcr 1-5* |
| 14 | Reddy et al., 2022[14] | 1852 | 31 | Telangana | VITEK-2, BMD | *mcr-1* |
| 15 | Das et al., 2022[15] | 158 | 27 | Odisha, UP, Rajasthan | BMD, VITEK-2 | *mcr 1-5* |
| 16 | Nirwan et al., 2021[16] | 6765 | 18 | Haryana | BMD, VITEK-2 | *mcr-1* |
| 17 | Azam et al., 2021[17] | 335 | 11 | Delhi | BMD, KB DDT | *mcr 1-8* |
| 18 | Kar et al., 2021[18] | 200 | 27 | Odisha | BMD, ADM, E test, RPNP | *mcr-1* & *mcr-2* |
| 19 | Sohail et al., 2021[19] | 106 | 19 | Karnataka | BMD, KB DDT | NO |
| 20 | Gunalan et al., 2021[20] | 75 | 11 | Pondicherry | BMD | NO |
| 21 | Priyanka et al., 2021[21] | 171 | 102 | Rajasthan | BMD | NO |
| 22 | Bandyopadhyay et al., 2021[22] | 72 | 2 | West Bengal | KB DDT, BMD | YES |
| 23 | Sharma et al., 2020[23] | 365 | 9 | Uttar Pradesh | BMD, KB DDT | *mcr 1-5* |
| 24 | Aarthi et al., 2020[24] | 440 | 11 | Tamil Nadu | BMD | YES |
| 25 | Raghupathi et al., 2020[25] | 30 | 10 | Tamil Nadu | BMD, KB DDT | *mcr 1-4* |
| 26 | Bardhan et al., 2020[26] | 84 | 57 | West Bengal | BMD, KB DDT | *mcr-1* |
| 27 | Khurana et al., 2020[27] | 910 | 196 | Delhi | VITEK-2, BMD | NO |
| 28 | Mitra et al., 2020[28] | 60 | 24 | Odisha | BMD | *mcr-1* & *mcr-2* |
| 29 | Das et al., 2020[29] | 138 | 31 | West Bengal | VITEK-2, BMD, E-test, Agar Diffusion | *mcr-1* |
| 30 | Waattal et al., 2020[30] | 225 | 73 | Delhi | Vitek-2, Micronaut-S, BMD E-test | NO |

| | | | | | | |
|---|---|---|---|---|---|---|
| 31 | Gogry et al., 2019[31] | 253 | 47 | Delhi | BMD | *mcr-1* & *mcr-3* |
| 32 | Mathur et al., 2019[32] | 846 | 34 | Delhi | VITEK-2, BMD, KB-DDT | YES |
| 33 | Sundaramoorthy et al., 2019 A[33] | 9 | 4 | Tamil Nadu | BMD | NO |
| 34 | Sundaramoorthy et al., 2019 B[34] | 6 | 2 | Tamil Nadu | BMD | *mcr 1-9* |
| 35 | Amladi et al.,2019[35] | 150 | 14 | Tamil Nadu | BMD | *mcr-1* |
| 36 | Garg et al.,2019[36] | 146 | 27 | Uttar Pradesh | BMD | NO |
| 37 | Raghupati et al.,2019[37] | 87 | 27 | Tamil Nadu | BMD, KB-DDT | NO |
| 38 | Kumar et al., 2018[38] | 932 | 17 | Kerala | VITEK-2, BMD | *mcr-1* & *mcr-2* |
| 39 | Manohar et al., 2017[39] | 89 | 29 | Tamil Nadu | KB DDT, BMD | *mcr-1* & *mcr-2* |
| 40 | Kumar., 2016[40] | 31 | 30 | Odisha & Haryana | BMD | NO |
| 41 | Kumar et al., 2016 C[41] | 1590 | 123 | Odisha | BMD, KB DDT | NO |

KB DDT- Kirby-Bauer Disc Diffusion Test, BMD- Broth Micro-dilution, Etest- Epsilometer Test, ADM- Agar Dilution Method, CBDE- Colistin Broth Disc Elution, RPNP- Rapid Polymyxin Nordmann Poirel Test. # The paper was published in 2023 as an online early version, hence included in the study.

**Supplementary Table 2**. Reports on the contribution of *mcr* gene in the development of polymyxin resistance in India.

| Sl. No. | Author(s) | State/ UT | No. of Pol$^R$ Isolates | No. of *mcr* positive isolates |
|---|---|---|---|---|
| 1 | #Seethalakshmi et al., 2024[42] | Tamil Nadu | 1 | 1 |
| 2 | Naha et al., 2023[43] | West Bengal | - | 10 |
| 3 | Premnath et al., 2023[44] | Tamil Nadu | - | 23 |
| 4 | Talat et al., 2023[45] | West Bengal | 1 | 1 |
| 5* | Pathak et al., 2023[46] | Uttar Pradesh | 6 | 0 |
| 6 | Aldeia et al., 2023[47] | Not mentioned | 1 | 0 |
| 7 | Sreejith et al., 2023[48] | Kerala | 1 | 1 |
| 8* | Shanthini et al., 2023[7] | Tamil Nadu | 27 | 0 |
| 9* | Rout et al., 2023[8] | Odisha | 778 | 0 |
| 10 | Talat et al., 2022[49] | West Bengal | 1 | 1 |
| 11* | Elizabeth et al., 2022[10] | Assam | 12 | 5 |
| 12* | Bir et al., 2022[11] | Delhi | 25 | 1 |
| 13* | Naha et al., 2022[50] | West Bengal | 9 | 0 |
| 14* | Sharma et al., 2022[13] | Uttar Pradesh | 25 | 0 |
| 15* | Reddy et al., 2022[14] | Telangana | 31 | 2 |
| 16* | Das et al., 2022[15] | Odisha, Uttar Pradesh, Rajasthan | 27 | 0 |
| 17* | Azam et al., 2021[17] | Delhi | 11 | 0 |
| 18* | Nirwan et al., 2021[16] | Haryana | 18 | 0 |
| 19* | Singh et al., 2021[51] | Uttar Pradesh | 22 | 19 |
| 20* | Elizabeth et al., 2021[52] | Assam | 6 | 2 |
| 21* | Kar et al., 2021[18] | Odisha | 27 | 0 |
| 22 | Karade et al., 2021[53] | Maharashtra | 1 | 0 |
| 23* | Bandyopadhyay et al., 2021[22] | West Bengal | 2 | 0 |
| 24* | Sharma et al., 2021[23] | Uttar Pradesh | 9 | 0 |
| 25* | Aarthi et al., 2021[24] | Tamil Nadu | 11 | 0 |
| 26* | Pathak et al., 2021[54] | Uttar Pradesh | 40 | 4 |
| 27* | Raghupathi et al., 2020[25] | Tamil Nadu | 10 | 1 |
| 28* | Roy et al., 2020[55] | West Bengal | 3 | 3 |
| 29 | Dey et al., 2020[56] | Odisha | 1 | 0 |
| 30* | Bardhan et al., 2020[26] | West Bengal | 57 | 0 |
| 31 | Sahoo et al., 2020[57] | Odisha | 1 | 0 |
| 32* | Khamari et al.,2020[58] | Andhra Pradesh | 2 | 0 |
| 33* | Palani et al., 2020[59] | Tamil Nadu | 43 | 2 |
| 34* | Mitra et al., 2020[28] | Odisha | 24 | 2 |
| 35 | Naha et al., 2020[60] | West Bengal | 1 | 0 |
| 36* | Das et al., 2020[29] | West Bengal | 31 | 0 |
| 37 | Pathak et al., 2020[61] | Uttar Pradesh | 1 | 1 |
| 38 | Bean et al., 2019[62] | West Bengal | 1 | 0 |
| 39* | Gogry et al., 2019[31] | Delhi | 47 | 5 |

| | | | | |
|---|---|---|---|---|
| 40* | Shankar et al., 2019[63] | Tamil Nadu | 65 | 0 |
| 41* | Mathur et al., 2019[32] | Delhi | 34 | 0 |
| 42 | Paul et al., 2019[64] | Kerala | 1 | 0 |
| 43* | Sundaramoorthy et al., 2019b[34] | Tamil Nadu | 2 | 0 |
| 44* | Subramaniam & Muthukrishnan, 2019[65] | Tamil Nadu | 2 | 2 |
| 45* | Amladi et al.,2019[35] | Tamil Nadu | 14 | 0 |
| 46* | Rahman & Ahmad., 2019[66] | Uttar Pradesh | 20 | 20 |
| 47* | Ghafur et al., 2019[67] | Tamil Nadu | 71 | 3 |
| 48* | Kumar et al., 2018[38] | Kerala | 17 | 0 |
| 49 | Shankar et al., 2018[68] | Tamil Nadu | 1 | 0 |
| 50* | Singh et al.,2018[69] | Uttar Pradesh | 21 | 4 |
| 51* | Aggarwal et al.,2018[70] | Delhi | 7 | 0 |
| 52* | Mathur et al., 2018[71] | Tamil Nadu | 8 | 0 |
| 53* | Manohar et al., 2017[39] | Tamil Nadu | 29 | 0 |
| 54* | Pragasam et al., 2017[72] | Tamil Nadu | 8 | 0 |
| 55 | Marathe et al., 2017[73] | Maharashtra | 1 | 1 |
| 56 | Veerarghavan et al., 2016[74] | Tamil Nadu | 1 | 0 |
| 57* | Bernasconi et al., 2016[75] | Not mentioned | 5 | 1 |
| 58 | Kumar et al., 2016 A[76] | Haryana | 1 | 1 |

### The paper was published in 2023 as an online early version, hence included in the study.
* indicates the papers used in the meta-analysis.

**Supplementary Table 3-** PRISMA 2020 Checklist.

| Section and Topic | Item# | Checklist item | Location where item is reported |
|---|---|---|---|
| **TITLE** | | | |
| Title | 1 | Identify the report as a systematic review. | 1 |
| **ABSTRACT** | | | |
| Abstract | 2 | See the PRISMA 2020 for Abstracts checklist. | 2 |
| **INTRODUCTION** | | | |
| Rationale | 3 | Describe the rationale for the review in the context of existing knowledge. | 3-5 |
| Objectives | 4 | Provide an explicit statement of the objective(s) or question(s) the review addresses. | 4-5 |
| **METHODS** | | | |
| Eligibility criteria | 5 | Specify the inclusion and exclusion criteria for the review and how studies were grouped for the syntheses. | 5-6 |
| Information sources | 6 | Specify all databases, registers, websites, organisations, reference lists and other sources searched or consulted to identify studies. Specify thedate when each source was last searched or consulted. | 5 |
| Search strategy | 7 | Present the full search strategies for all databases, registers and websites, including any filters and limits used. | 5 |
| Selection process | 8 | Specify the methods used to decide whether a study met the inclusion criteria of the review, including how many reviewers screened each recordand each report retrieved, whether they worked independently, and if applicable, details of automation tools used in the process. | 5-6 |
| Data collection process | 9 | Specify the methods used to collect data from reports, including how many reviewers collected data from each report, whether they worked independently, any processes for obtaining or confirming data from study investigators, and if applicable, details of automation tools used in theprocess. | 6 |
| Data items | 10a | List and define all outcomes for which data were sought. Specify whether all results that were compatible with each outcome domain in each study were sought (e.g. for all measures, time points, analyses), and if not, the methods used to decide which results to collect. | 6 |
| | 10b | List and define all other variables for which data were sought (e.g. participant and intervention characteristics, funding sources). Describe any assumptions made about any missing or unclear information. | 6-7 |
| Study risk of bias assessment | 11 | Specify the methods used to assess risk of bias in the included studies, including details of the tool(s) used, how many reviewers assessed each study and whether they worked independently, and if applicable, details of automation tools used in the process. | 6 |
| Effect measures | 12 | Specify for each outcome the effect measure(s) (e.g. risk ratio, mean difference) used in the synthesis or presentation of results. | 6 |
| Synthesis methods | 13a | Describe the processes used to decide which studies were eligible for each synthesis (e.g. tabulating the study intervention | N/A |

| | | | |
|---|---|---|---|
| | | characteristics andcomparing against the planned groups for each synthesis (item #5)). | |
| | 13b | Describe any methods required to prepare the data for presentation or synthesis, such as handling of missing summary statistics, or data conversions. | 6 |
| | 13c | Describe any methods used to tabulate or visually display results of individual studies and syntheses. | 6 |
| | 13d | Describe any methods used to synthesize results and provide a rationale for the choice(s). If meta-analysis was performed, describe the model(s), method(s) to identify the presence and extent of statistical heterogeneity, and software package(s) used. | 6-7 |
| | 13e | Describe any methods used to explore possible causes of heterogeneity among study results (e.g. subgroup analysis, meta-regression). | 6-7 |
| | 13f | Describe any sensitivity analyses conducted to assess robustness of the synthesized results. | 7 |
| Reporting bias assessment | 14 | Describe any methods used to assess risk of bias due to missing results in a synthesis (arising from reporting biases). | 7 |
| Certainty assessment | 15 | Describe any methods used to assess certainty (or confidence) in the body of evidence for an outcome. | 7 |
| **Section andTopic** | **Item#** | **Checklist item** | **Location where item is reported** |
| **RESULTS** | | | |
| Study selection | 16a | Describe the results of the search and selection process, from the number of records identified in the search to the number of studies included inthe review, ideally using a flow diagram. | 7 |
| | 16b | Cite studies that might appear to meet the inclusion criteria, but which were excluded, and explain why they were excluded. | 7 |
| Study characteristics | 17 | Cite each included study and present its characteristics. | Suppl. Info. |
| Risk of bias instudies | 18 | Present assessments of risk of bias for each included study. | 7, Table-1 |
| Results of individual studies | 19 | For all outcomes, present, for each study: (a) summary statistics for each group (where appropriate) and (b) an effect estimate and its precision(e.g. confidence/credible interval), ideally using structured tables or plots. | Fig. 1, Table-1 |
| Results of syntheses | 20a | For each synthesis, briefly summarise the characteristics and risk of bias among contributing studies. | Table-1, Suppl. Fig. 1& 2 |
| | 20b | Present results of all statistical syntheses conducted. If meta-analysis was done, present for each the summary estimate and its precision (e.g.confidence/credible interval) and measures of statistical heterogeneity. If comparing groups, describe the direction of the effect. | Table-1 |
| | 20c | Present results of all investigations of possible causes of heterogeneity among study results. | 8, Table-1 |
| | 20d | Present results of all sensitivity analyses conducted to assess the robustness of the synthesized results. | 8, Suppl. Fig. 3 & 4 |

| | | | |
|---|---|---|---|
| Reporting biases | 21 | Present assessments of risk of bias due to missing results (arising from reporting biases) for each synthesis assessed. | ---- |
| Certainty of evidence | 22 | Present assessments of certainty (or confidence) in the body of evidence for each outcome assessed. | ---- |
| **DISCUSSION** | | | |
| Discussion | 23a | Provide a general interpretation of the results in the context of other evidence. | 8-11 |
| | 23b | Discuss any limitations of the evidence included in the review. | 11 |
| | 23c | Discuss any limitations of the review processes used. | 11 |
| | 23d | Discuss implications of the results for practice, policy, and future research. | 11 |
| **OTHER INFORMATION** | | | |
| Registration and protocol | 24a | Provide registration information for the review, including register name and registration number, or state that the review was not registered. | N/A |
| | 24b | Indicate where the review protocol can be accessed, or state that a protocol was not prepared. | 5-6 |
|  | 24c | Describe and explain any amendments to information provided at registration or in the protocol. | ---- |
| Support | 25 | Describe sources of financial or non-financial support for the review, and the role of the funders or sponsors in the review. | 12 |
| Competing interests | 26 | Declare any competing interests of review authors. | 12 |
| Availability of data, code and other materials | 27 | Report which of the following are publicly available and where they can be found: template data collection forms; data extracted from included studies; data used for all analyses; analytic code; any other materials used in the review. | Suppl. Information |

*From:* Page MJ, McKenzie JE, Bossuyt PM, Boutron I, Hoffmann TC, Mulrow CD, et al. The PRISMA 2020 statement: an updated guideline for reporting systematic reviews. BMJ 2021;372:n71. doi: 10.1136/bmj.n71

For more information, visit: http://www.prisma-statement.org/

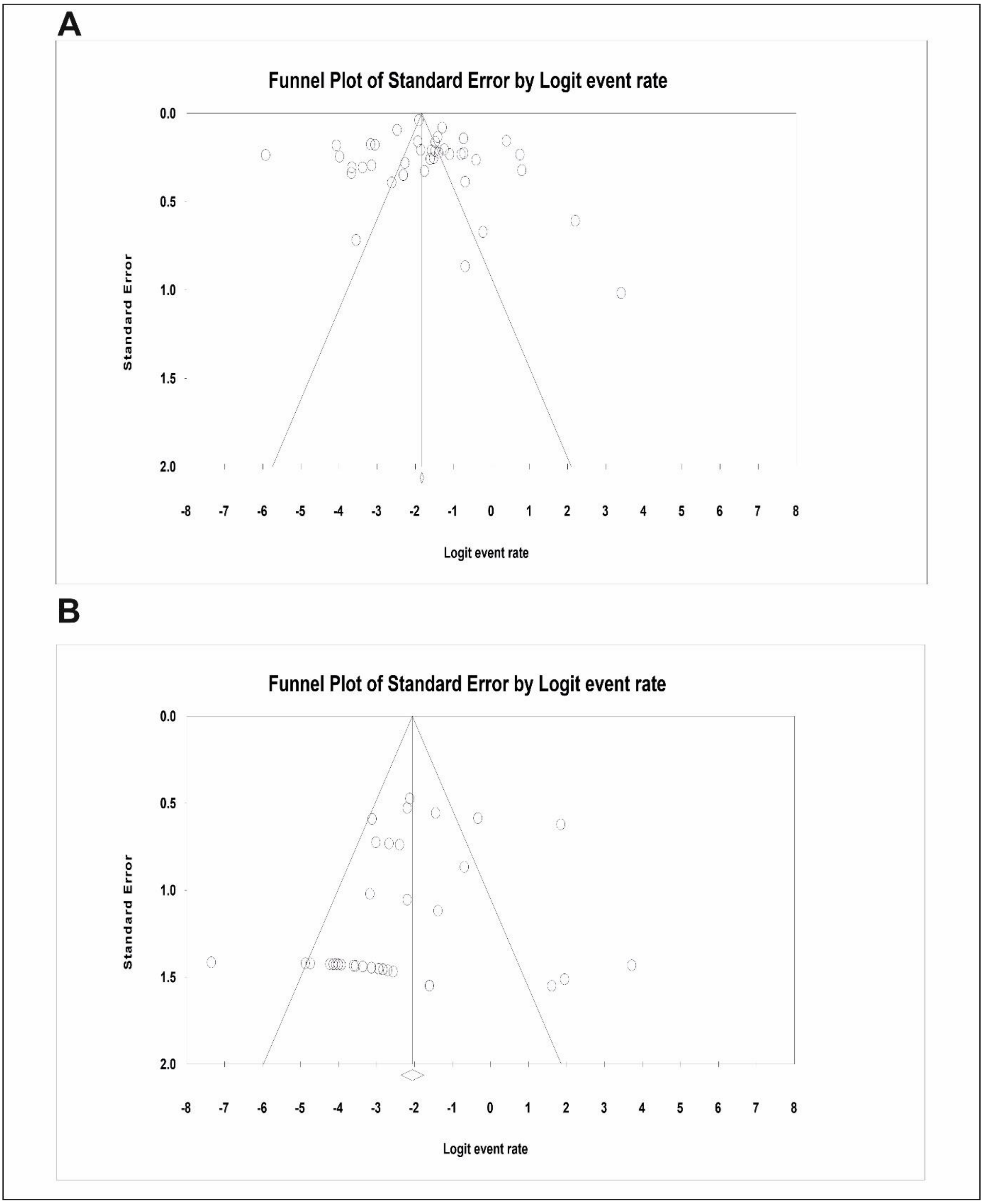


**Supplementary Figure 1.** Publication bias determination by Funnel plot of polymyxin resistant bacteria in India (A) and contribution of *mcr* gene in the development of polymyxin resistance (B).

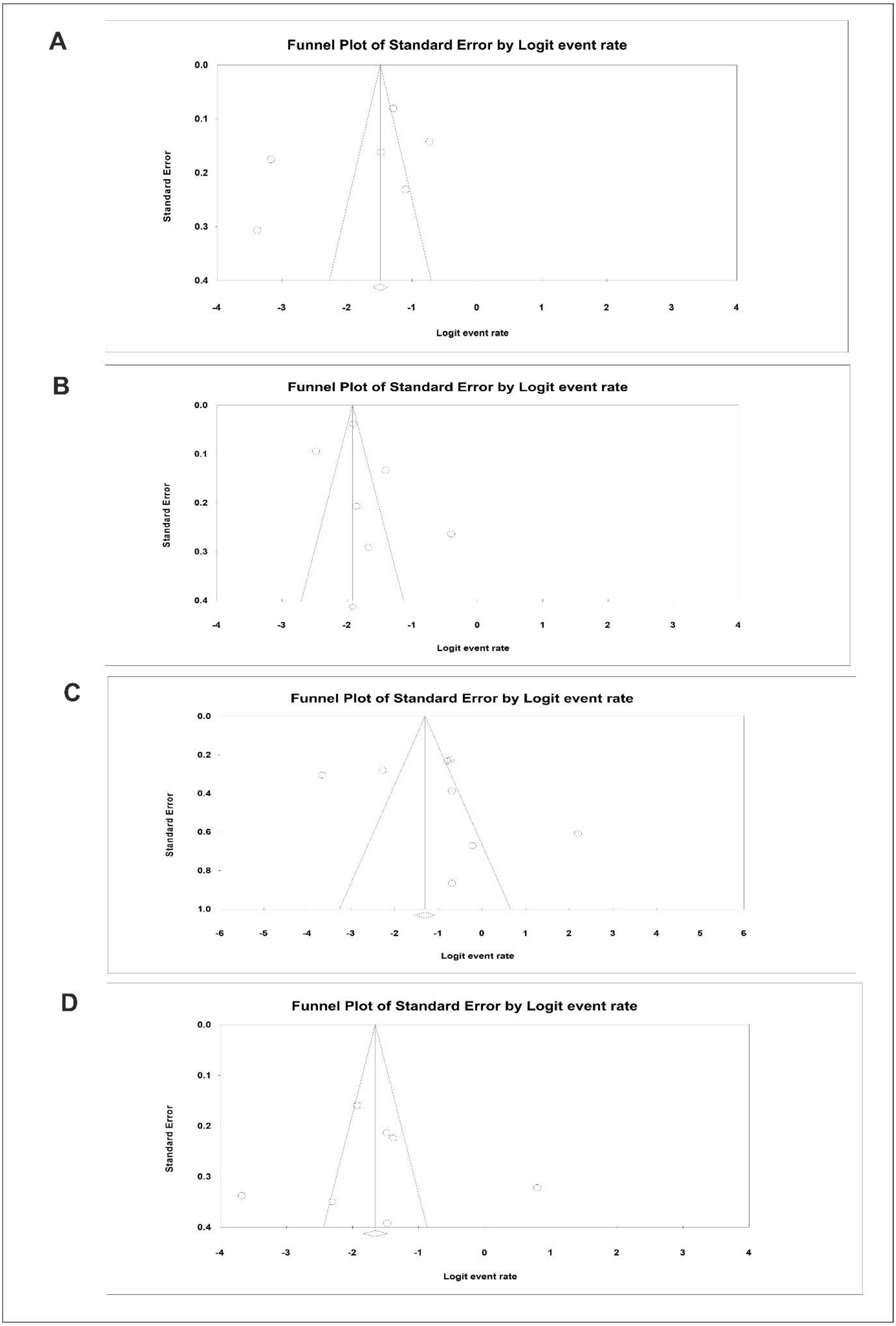
A
Funnel Plot of Standard Error by Logit event rate
Standard Error
0.0
0.1
0.2
0.3
0.4
-4
-3
-2
-1
0
1
2
3
4
Logit event rate
B
Funnel Plot of Standard Error by Logit event rate
Standard Error
0.0
0.1
0.2
0.3
0.4
-4
-3
-2
-1
0
1
2
3
4
Logit event rate
C
Funnel Plot of Standard Error by Logit event rate
Standard Error
0.0
0.2
0.4
0.6
0.8
1.0
-6
-5
-4
-3
-2
-1
0
1
2
3
4
5
6
Logit event rate
D
Funnel Plot of Standard Error by Logit event rate
Standard Error
0.0
0.1
0.2
0.3
0.4
-4
-3
-2
-1
0
1
2
3
4
Logit event rate

**Supplementary Figure 2.** Publication bias determination by Funnel plot of polymyxin-resistant bacteria in different states of India, (A) Delhi, (B) Odisha, (C) Tamil Nadu, and (D) Uttar Pradesh.

| Study name | Statistics with study removed | | | | Event rate (95% CI) with study removed |
|---|---|---|---|---|---|
| | Point | Lower limit | Upper limit | Z-Value | |
| Baneerjee et al., 2024 | 0.152 | 0.113 | 0.201 | -9.841 | |
| Vasesi et al., 2024 | 0.153 | 0.113 | 0.202 | -9.819 | |
| Soni et al., 2023 | 0.154 | 0.115 | 0.204 | -9.766 | |
| Kaza et al., 2023 | 0.149 | 0.111 | 0.199 | -9.902 | |
| Sharma et al., 2023 | 0.151 | 0.111 | 0.201 | -9.743 | |
| Ranjan et al., 2023 | 0.152 | 0.113 | 0.201 | -9.841 | |
| Shanthini et al., 2023 | 0.140 | 0.104 | 0.185 | -10.575 | |
| Rout et al., 2023 | 0.153 | 0.107 | 0.213 | -8.233 | |
| Sharma et al., 2022 A | 0.142 | 0.106 | 0.188 | -10.505 | |
| Elizabeth et al., 2022 | 0.154 | 0.115 | 0.204 | -9.760 | |
| Bir et al., 2022 | 0.148 | 0.109 | 0.196 | -9.992 | |
| Panigrahi et al., 2022 | 0.149 | 0.110 | 0.199 | -9.780 | |
| Sharma et al, 2022 B | 0.149 | 0.110 | 0.198 | -9.923 | |
| Reddy et al., 2022 | 0.157 | 0.118 | 0.206 | -9.991 | |
| Das et al., 2022 | 0.149 | 0.111 | 0.199 | -9.875 | |
| Nirwan et al., 2021 | 0.163 | 0.125 | 0.210 | -10.348 | |
| Azam et al., 2021 | 0.155 | 0.116 | 0.205 | -9.750 | |
| Kar et al., 2021 | 0.150 | 0.111 | 0.200 | -9.824 | |
| Sohail et al., 2021 | 0.149 | 0.111 | 0.198 | -9.915 | |
| Gunalan et al., 2021 | 0.150 | 0.111 | 0.199 | -9.906 | |
| Priyanka et al., 2021 | 0.142 | 0.107 | 0.186 | -10.860 | |
| Bandyopadhyay et al., 2021 | 0.154 | 0.115 | 0.204 | -9.798 | |
| Aarthi et al., 2020 | 0.156 | 0.116 | 0.206 | -9.743 | |
| Raghupati et al., 2020 | 0.147 | 0.109 | 0.195 | -10.098 | |
| Sharma et al., 2020 | 0.156 | 0.116 | 0.206 | -9.736 | |
| Bardhan et al., 2020 | 0.141 | 0.106 | 0.186 | -10.678 | |
| Khurana et al., 2020 | 0.149 | 0.109 | 0.200 | -9.512 | |
| Waattal et al., 2020 | 0.146 | 0.109 | 0.195 | -10.085 | |
| Mitra et al., 2020 | 0.145 | 0.108 | 0.193 | -10.179 | |
| Das et al., 2020 | 0.148 | 0.110 | 0.197 | -9.942 | |
| Gogry et al., 2019 | 0.149 | 0.110 | 0.199 | -9.831 | |
| Mathur et al., 2019 | 0.155 | 0.115 | 0.205 | -9.782 | |
| Sundaramoorthy et al., 2019 A | 0.146 | 0.108 | 0.194 | -10.156 | |
| Sundaramoorthy et al., 2019 B | 0.148 | 0.110 | 0.196 | -10.091 | |
| Amladi et al., 2019 | 0.152 | 0.112 | 0.201 | -9.822 | |
| Garg et al., 2019 | 0.149 | 0.110 | 0.198 | -9.895 | |
| Raghupati et al., 2019 | 0.147 | 0.109 | 0.195 | -10.070 | |
| Kumar et al., 2018 | 0.157 | 0.118 | 0.207 | -9.799 | |
| Manohar et al., 2017 | 0.146 | 0.109 | 0.195 | -10.090 | |
| Kumar et al., 2016 A | 0.141 | 0.105 | 0.187 | -10.505 | |
| Kumar et al., 2016 B | 0.153 | 0.112 | 0.204 | -9.513 | |
| | 0.150 | 0.112 | 0.198 | -10.078 | |

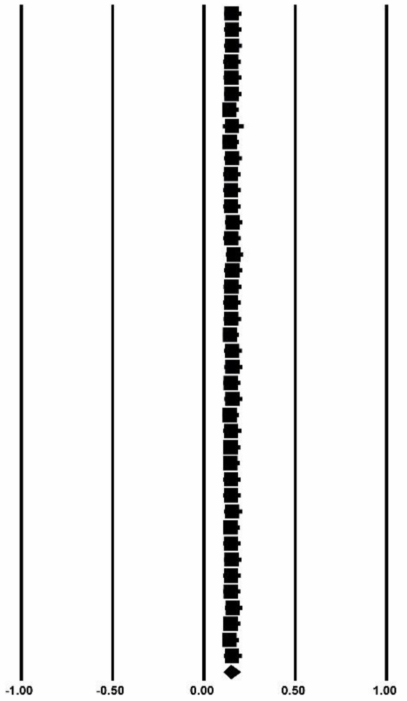


**Supplementary Figure 3**. Sensitivity plot demonstrating the incidence of polymyxin-resistant bacteria in India.

| Study name | Point | Lower limit | Upper limit | Z-Value |
|---|---|---|---|---|
| | Statistics with study removed | | | |
| Pathak et al., 2023 | 0.084 | 0.048 | 0.145 | -7.690 |
| Shanthini et al., 2023 | 0.087 | 0.049 | 0.148 | -7.627 |
| Rout et al., 2023 | 0.094 | 0.055 | 0.156 | -7.701 |
| Elizabeth et al., 2022 | 0.079 | 0.044 | 0.135 | -7.989 |
| Bir et al., 2022 | 0.086 | 0.048 | 0.147 | -7.601 |
| Naha et al., 2022 | 0.085 | 0.048 | 0.146 | -7.668 |
| Sharma et al, 2022 | 0.087 | 0.049 | 0.148 | -7.629 |
| Reddy et al., 2022 | 0.084 | 0.047 | 0.146 | -7.560 |
| Das et al., 2022 | 0.087 | 0.049 | 0.148 | -7.627 |
| Azam et al., 2021 | 0.085 | 0.048 | 0.146 | -7.659 |
| Nirwan et al., 2021 | 0.086 | 0.049 | 0.147 | -7.639 |
| Singh et al., 2021 | 0.075 | 0.046 | 0.120 | -9.479 |
| Elizabeth et al., 2021 | 0.080 | 0.045 | 0.138 | -7.848 |
| Kar et al., 2021 | 0.087 | 0.049 | 0.148 | -7.627 |
| Bandyopadhyay et al., 2021 | 0.083 | 0.047 | 0.142 | -7.760 |
| Pathak et al., 2021 | 0.083 | 0.046 | 0.145 | -7.492 |
| Aarthi et al., 2020 | 0.085 | 0.048 | 0.146 | -7.659 |
| Raghupati et al., 2020 | 0.083 | 0.047 | 0.144 | -7.667 |
| Roy et al., 2020 | 0.078 | 0.045 | 0.132 | -8.181 |
| Sharma et al., 2020 | 0.085 | 0.048 | 0.146 | -7.668 |
| Bardhan et al., 2020 | 0.088 | 0.050 | 0.150 | -7.616 |
| Khamari et al., 2020 | 0.083 | 0.047 | 0.142 | -7.760 |
| Palani et al., 2020 | 0.085 | 0.048 | 0.147 | -7.547 |
| Mitra et al., 2020 | 0.084 | 0.047 | 0.145 | -7.580 |
| Das et al., 2020 | 0.087 | 0.050 | 0.149 | -7.624 |
| Gogry et al., 2019 | 0.083 | 0.046 | 0.145 | -7.452 |
| Shankar et al., 2019 | 0.089 | 0.051 | 0.150 | -7.615 |
| Mathur et al., 2019 | 0.087 | 0.050 | 0.149 | -7.622 |
| Sundaramoorthy et al., 2019 B | 0.083 | 0.047 | 0.142 | -7.760 |
| Subramaniam & Muthukrishnan, 2019 | 0.078 | 0.045 | 0.134 | -8.116 |
| Amladi et al., 2019 | 0.086 | 0.049 | 0.147 | -7.648 |
| Rahman & Ahmad, 2019 | 0.076 | 0.044 | 0.126 | -8.587 |
| Kumar et al., 2018 | 0.086 | 0.049 | 0.147 | -7.641 |
| Ghafur et al., 2018 | 0.086 | 0.048 | 0.148 | -7.510 |
| Singh et al., 2018 | 0.081 | 0.045 | 0.141 | -7.629 |
| Aggarwal et al., 2018 | 0.084 | 0.048 | 0.145 | -7.682 |
| Mathur et al., 2018 | 0.085 | 0.048 | 0.145 | -7.675 |
| Manohar et al., 2017 | 0.087 | 0.049 | 0.148 | -7.625 |
| Pragasam et al., 2017 | 0.085 | 0.048 | 0.145 | -7.675 |
| Bernasconi et al., 2016 | 0.082 | 0.046 | 0.141 | -7.756 |
| | 0.084 | 0.048 | 0.143 | -7.835 |

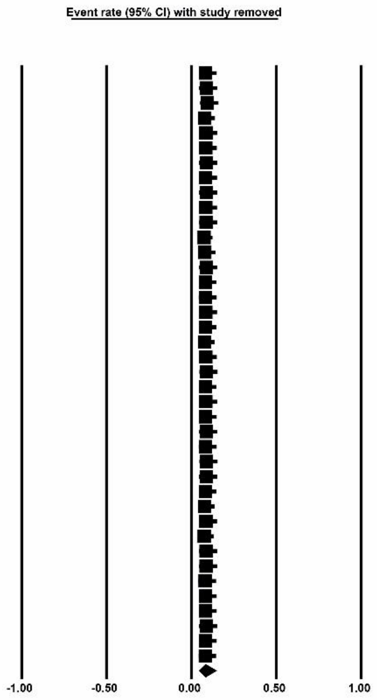


**Supplementary Figure 4**. Sensitivity plot demonstrating the contribution of the *mcr* gene in the development of polymyxin resistance in India.
